# Nanoparticle-Supported, Point-of-Care Detection of Shiga Toxin-Producing E. coli Infection from Food and Human Specimens

**DOI:** 10.1101/2025.04.03.25325182

**Authors:** Seyedsina Mirjalili, Yeji Choi, Karuppiah Chockalingam, Benjamin Thomas, Xiaohua He, Zhilei Chen, Chao Wang

## Abstract

Shiga toxin-producing *Escherichia coli* (STEC) are major foodborne pathogens responsible for severe infections, including the deadly hemolytic uremic syndrome (HUS). However, the current diagnostic methods lack the sensitivity and speed required for effective clinical and food safety applications. Early detection of Shiga toxin 2 (Stx2), a primary virulence factor of STEC, could potentially offer critical benefits for timely intervention. In this work, gold <u>n</u>anoparticles (AuNPs) are functionalized with a pair of high-affinity, <u>d</u>esigned <u>a</u>nkyrin repeat <u>p</u>roteins (DARPins) targeting the A and B subunits of Stx2, and used as multifunctional signal transductors for <u>r</u>apid and <u>e</u>lectronic <u>d</u>etection (RED). This DARPin-RED platform leverages active centrifugal forces and vortex agitation for signal enhancement within a short turnaround time (<30 minutes), achieving highly sensitive (attomolar to femtomolar) detection of Stx2 spiked in food matrices, such as milk, lettuce extract, and ground beef extract, as well as biological fluids, including whole blood, and serum. Additionally, DARPin-RED is capable of detecting multiple Stx2 subtypes without serious background interference, and successful in both differentiating high-toxin-producing E. coli strain (RM5856) from low toxin producer (RM9872) (p < 0.001) and analyzing different bacterial inoculation stages (p = 0.011) from STEC culture within 8 hours post-inoculation. The ability of DARPin-RED to detect Stx2 from food and human specimens at a high sensitivity and specificity using a point-of-care (POC) readout circuit presents a significant advancement for mitigating foodborne outbreaks and effective management of HUS progression.

## Introduction

Shiga toxin-producing *E. coli* (STEC) are zoonotic, pathogenic bacteria characterized by Shiga toxin production. STEC infection occurs following the ingestion of contaminated food or water or by direct contact with infected animals or humans^1^. STEC pose a significant public health concern due to their propensity to cause outbreaks and sporadic cases of bloody diarrhea. The Centers for Disease Control and Prevention (CDC) estimated that ∼265,000 STEC infections occur each year in the United States, causing 3,600 hospitalizations and 30 deaths^2^.

Patients typically develop diarrhea within about a week after STEC infection (e.g., E. coli strain O157:H7)^3^, and may progress with bloody diarrhea 3 to 5 days later^4^. A significant amount (up to 15%) of these STEC-infected patients will develop hemolytic uremic syndrome (HUS) in another few days^1,5^, leaving a small window for diagnosis and treatment. Importantly, antibiotics are found to induce the production of Stx2 phage and release of Stx2, thus increasing risks of developing HUS and even deaths^6–9^. HUS is characterized by acute renal failure, thrombocytopenia, and microangiopathic hemolytic anemia^10–12^, and has a mortality rate of 5-20%^13,14^. Among those that survive STEC-HUS, around 30% develop long-term sequelae after acute onset, including renal sequelae (proteinuria, chronic kidney disease, etc.) and neurological complications (tetraplegia, cognitive impairment, etc.)^15,16^. Thus, a fast and accurate diagnosis is necessary for patients presenting STEC-like symptoms prior to treatment^17^. On the other hand, STEC can survive and persist in numerous environments, such as farm soil, water, and food as well as in animal reservoirs^18^, resulting in frequent STEC outbreaks. Such outbreaks have been traced to the consumption of bacterial-contaminated food, such as beef^19^, diary products^20^, fresh vegetables^21^, etc. As one example, the consumption of contaminated sprouts was identified as the most likely cause of the 2011 STEC outbreak in Germany, affecting 3816 patients and causing 54 deaths. Among these, 845 patients developed HUS^22^.

The pathology of STEC stems from two exotoxins – Shiga toxin 1 (Stx1) and Shiga toxin 2 (Stx2), with Stx2 being the primary virulence factor^23^. Shiga toxins are extremely toxic, with a 50% lethal dose for mice estimated to be 50 ng/kg (∼0.7 pmol/kg)^24^. Additionally, the infection dose of STEC in the food matrices may be as low as 50 bacteria^25^, and the toxin concentration is also low in patient samples (<10 ng/mL in serum^26^). Therefore, a highly sensitive STEC test from both food matrices and patient samples is of particular importance. The conventional strategy for STEC diagnosis relies on the unique features of E. coli O157:H7 strains, such as delayed D-sorbitol fermentation (>24 h) and inability to produce β-glucuronidase^27^. However, non-O157 serotypes have been responsible for 113,000 illnesses annually in the United States alone^28^. Therefore, accurate STEC diagnosis requires the detection of either toxins or toxin-encoding genes.

For toxin detection, liquid chromatography-mass spectrometry (LC-MS) proved a limit-of-detection (LoD) of ∼120 pg/mL (1.7 pM) of Stx2^24^, Enzyme-Linked Immunosorbent Assay (ELISA) demonstrated a LoD of 25 pg/mL (357 fM) in ∼2.5 hours^29^, and cell culture-based assays using Hela or Vero cells could detect Stx2 with a LoD of 10-23 pg/mL (142-328 fM)^30,31^ but required 2-3 days of culture time. In comparison, Nucleic Acid Amplification Tests (NAATs) based methods, e.g. polymerase chain reaction (PCR) or loop-mediated-isothermal-amplification (LAMP), can achieve a very high detection sensitivity of the toxin-encoding gene (<100 CFU/mL)^32^. However, LC-MS, ELISA, or PCR-based STEC assays typically require execution in centralized labs and the culturing of stool specimens in enriched broth for 18-28 hours before detection^33^, causing significant delays in STEC diagnosis. Additionally, NAAT tests require extensive sample preparation to concentrate bacteria and extract DNA molecules for diagnosis. Their accuracies are complicated by the sample complexity and variability, often leading to false-negative diagnoses due to the presence of inhibiting enzymes and false-positive results due to mutations in target gene sequences (such as defective *stx* genes) in bacteria.

Rapid and accurate STEC diagnosis is desired not only for field use for contamination inspections to enhance food safety but also urgently needed for initiating timely and precise treatments for patients at risk. For example, due to the typical long time to return STEC test results, it is suggested that aggressive volume expansion should be initiated on all children suspected of STEC infection while the diagnosis is pending, in order to reduce the risks of HUS development^5^. Yet, since many other pathogens, such as *Chloridioides difficile* and *Salmonella typhimurim*, can also cause similar symptoms, including bloody diarrhea, treatment without conclusive STEC diagnosis inevitably increases risks for mistreating other infections and the medical cost.

In this study, we have developed a point-of-care (POC) diagnostic assay for detecting Stx2 in whole blood, and sera samples as well as food samples (lettuce, milk, and meat) with high (attomolar to femtomolar) sensitivity. Built upon two recently developed technologies, nanoparticle-supported, rapid, electronic detection (NasRED) assay^34–37^ and designed ankyrin repeat proteins (DARPins), our new platform, termed DARPin-RED, is designed to detect Stx2 using a pair of DARPins that target the A subunit and pentameric B subunit^38,39^, respectively. Our previous studies^31^ show that DARPin SHT (D_SHT_) targets the A-subunit of Stx2 and inhibits the toxin catalytic activity, and DARPin #3 (D_#3_) binds the basal surface of the B-subunit and is presumed to inhibit toxin attachment. In DARPin-RED, one set of gold nanoparticles (AuNPs) were functionalized with either mono-biotinylated D_SHT_ or D_#3_, and these AuNPs were mixed with samples of interest in a single tube. The AuNPs transduce the specific binding of DARPins to Stx2 into the physical aggregation of AuNPs and subsequent change in solution colorimetric signal. The optical signal is then digitized using an inexpensive portable electronic detector (PED) comprised of a light-emitting diode, a photodetector, and customized semiconductor-based circuitry with greatly improved stability^37^. Unlike other lab-based tests that require complex sample preparation, including purification, DNA extraction, washing, labeling, or enzymatic reactions, DARPin-RED achieved specific detection of Stx2 in diverse biological samples. This technological advancement would facilitate early detection and intervention, ensuring food security and improving patient outcomes.

## Results and discussion

### DARPin-RED platform

DARPin-RED as a POC setup assay was designed to reduce complexity, turnaround time, manual manipulation, and assay cost, as well as to increase the assay sensitivity, specificity, and accessibility. To achieve these, the platform leverages a pair of DARPin molecules previously engineered to bind specifically to Stx2^31^ and NasRED technology^34,35,37,40^. Firstly, the DARPins (SHT and #3) were mono-biotinylated and mounted on the surface of streptavidin-AuNPs to form a co-binder sensing solution specific for Stx2 toxin (**Fig. 1 a**). The sensing solution was then mixed with serially diluted Stx2a in a microcentrifuge tube to test the sensor performance. Upon mixing, Stx2a in the mixture binds the D_SHT_ and D_#3_ attached to different AuNPs, leading to reduced interparticle distances between these AuNPs and the formation of AuNP physical clusters. Higher Stx2a concentrations in solution correlate to larger cluster mass and more pronounced AuNP gravitational precipitation, which is manifested as a solution color change from reddishness (free-floating AuNPs absorption of green light at ∼570 nm) to transparent (medium without AuNPs) (**Fig. 1 b**).

**Figure 1.**
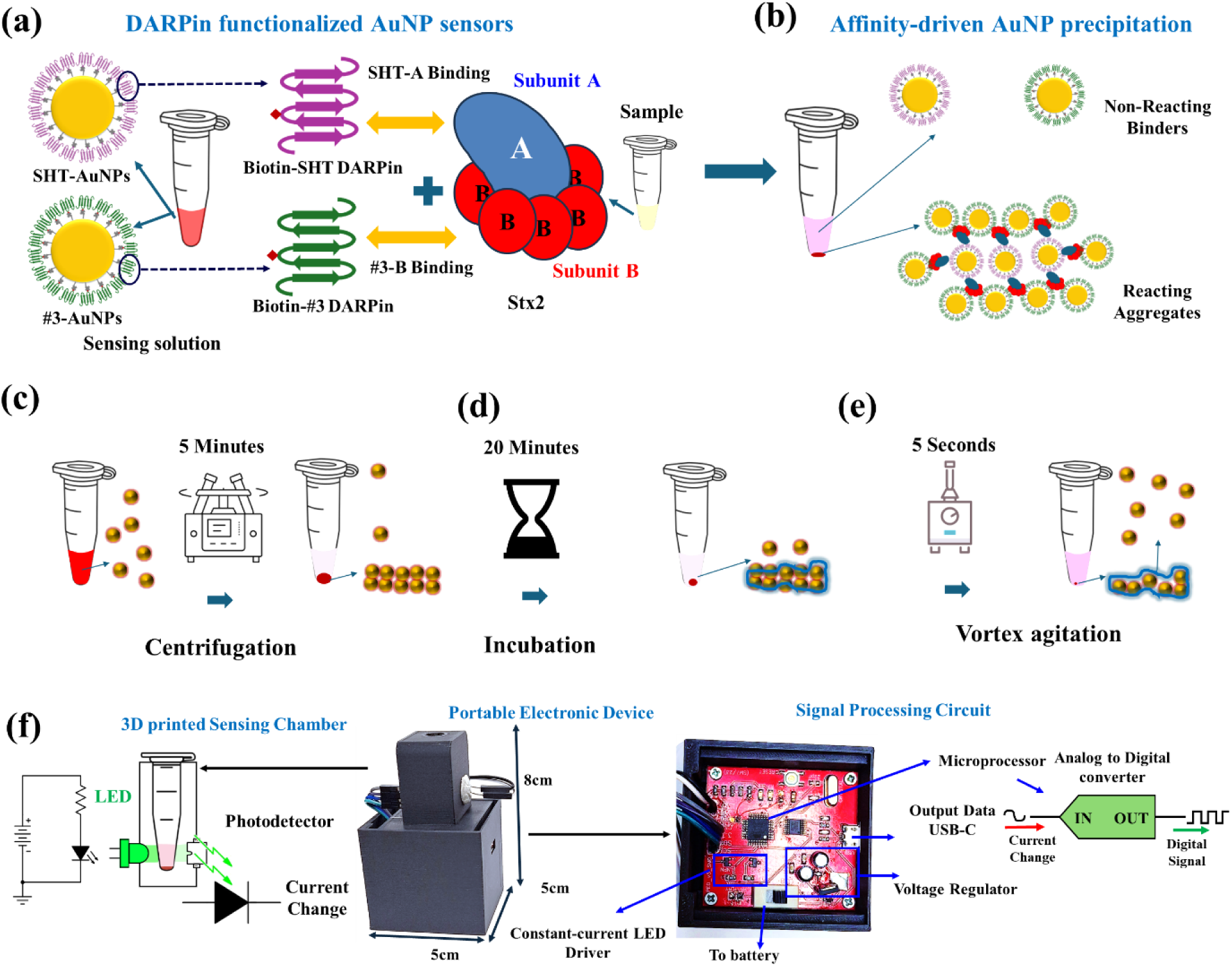
DARPin-RED design scheme. (a) Probe functionalization utilizing biotinylated DARPins and streptavidin covalent conjugated gold nanoparticles. Co-binders (SHT-AuNPs and #3-AuNps) mixing with titration of a spiked or authentic sample. (b) The coagulation of the co-binders in the presence of the Stx2 leads to precipitation of the clusters. (c) Accelerated reaction utilizing centrifugation to precipitate the AuNPs at the bottom of the tubes, enhancing the sensitivity and reducing reaction time, followed by (d) incubation to allow the cluster growth. (e) Vortex agitation is used to resuspend the non-reacting binders and preserve the high specificity of the assay. (f) the supernatant optical transmission readout using portable electronic device comprising a tube chamber, an LED and a photodetector. The changes in optical transmission are correlated with the Stx2 concentration and will lead to a change in the photodetector current. The custom circuitry is comprised of a constant current LED driver to stabilize the emitted light intensity of the LED, a voltage regulator feedback loop including a Zener diode to reduce and stabilize photodetector signals, and a microcontroller to convert the current changes into digital signals and mediate the connection between the device and a laptop.

The clusterization process typically takes 3 to 5 hours without intervention due to the slow passive diffusion and sedimentation rate. Similar assay time is seen in many diagnostics methods (i.e., ELISA) that rely on passive diffusion of the target in the medium^35^ and require extended incubation time to achieve diffusion equilibrium. To accelerate the detection process, the NasRED platform introduces an innovative and simple protocol that uses a benchtop centrifuge and a vortex unit to modify the reaction parameters. The sedimentation time of AuNPs in NasRED follows the equation 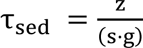, where z is the precipitation length, g is the gravitation constant, and s is the sedimentation coefficient determined by AuNP diameter, buffer density, and buffer viscosity. To accelerate the clusterization process, the solution is centrifuged (increasing g) to increase the collision rate of the target with functionalized AuNPs. This centrifugation step also creates a pellet at the bottom of the tube (decreasing z) with high AuNP concentration (**Fig. 1 c**), which promotes rapid growth of stable AuNP clusters catalyzed by Stx2a bridging different DARPin-AuNPs, leading to the formation of stable clusters within 20 minutes (**Fig. 1 d**). The entire mixtures are then subjected to an engineered fluidic force (i.e., 1950 RPM vortex agitation), which creates a turbulent flow and redisperses all the non-reacting, free AuNPs but not the clustered AuNPs due to their larger mass and the drag force^37^ (**Fig. 1 e**).

The Stx2a concentration in solution is directly correlated with the size of clusters, which affects the optical transmission of the resulting supernatant comprised of the non-reacting AuNPs. Such optical signals are then digitized using our custom PED, which harbors a 3D printed carbon filament tube to cancel the ambient light noise, an LED to emit the light on the supernatant of the tube at a wavelength of maximum AuNPs adsorption (∼570 nm for 80 nm AuNPs), a photodetector (PD) that converts the transmitted light to changes in current, and an inexpensive customized semiconductor circuitry integrated with a microprocessor to stabilize both the LED emission and PD signal and digitize the current changes (**Fig. 1 f**). The digital signals are subsequently transmitted to a computer using USB, Wi-Fi, or Bluetooth modules and analyzed using Python scripts for NasRED signal processing (see methods).

### DARPin-RED sensor performance enhancement by targeting distant sites on Stx2

The geometry of the binders-Stx2a complex impacts the sensitivity of the NasRED assay for Stx2a, and the selection of different targeting epitopes on Stx2a can affect the ratio of precipitated AuNPs to Stx2a molecules in solution. As demonstrated in previous studies, targeting distant epitopes with differentially functionalized AuNPs is an effective strategy to achieve this goal^35,37^(**Fig. 2a**). To investigate this effect, we first evaluated mono-binders – SHT-AuNPs (targeting the A-subunit), #3-AuNPs (targeting B subunit), and G1-AuNPs (nanobody targeting the B-subunit^41^) – against serially diluted Stx2a in PBSD (1xPBS, 20% (v/v) glycerol and 1 wt% BSA). In these experiments, 6 µL diluted Stx2a was mixed with 18 µL functionalized-AuNP (resuspended in PBSD) in each microcentrifuge tube, and the mixture was subjected to the mono-cycle NaSRED protocol (see methods). As expected, SHT-AuNPs alone did not induce AuNP clusterization, as they bind exclusively to the A-subunit without facilitating interparticle bridging, resulting in no detectable signal (**Fig. 2 b**). In contrast, 3-AuNPs and G1-AuNPs successfully detected Stx2a which is attributed to the pentameric architecture of the Stx2 B-subunit, which provides up to five distinct binding sites for AuNP attachment (**Fig. 2 a, c-e**, **Table 1**).

**Figure 2.**
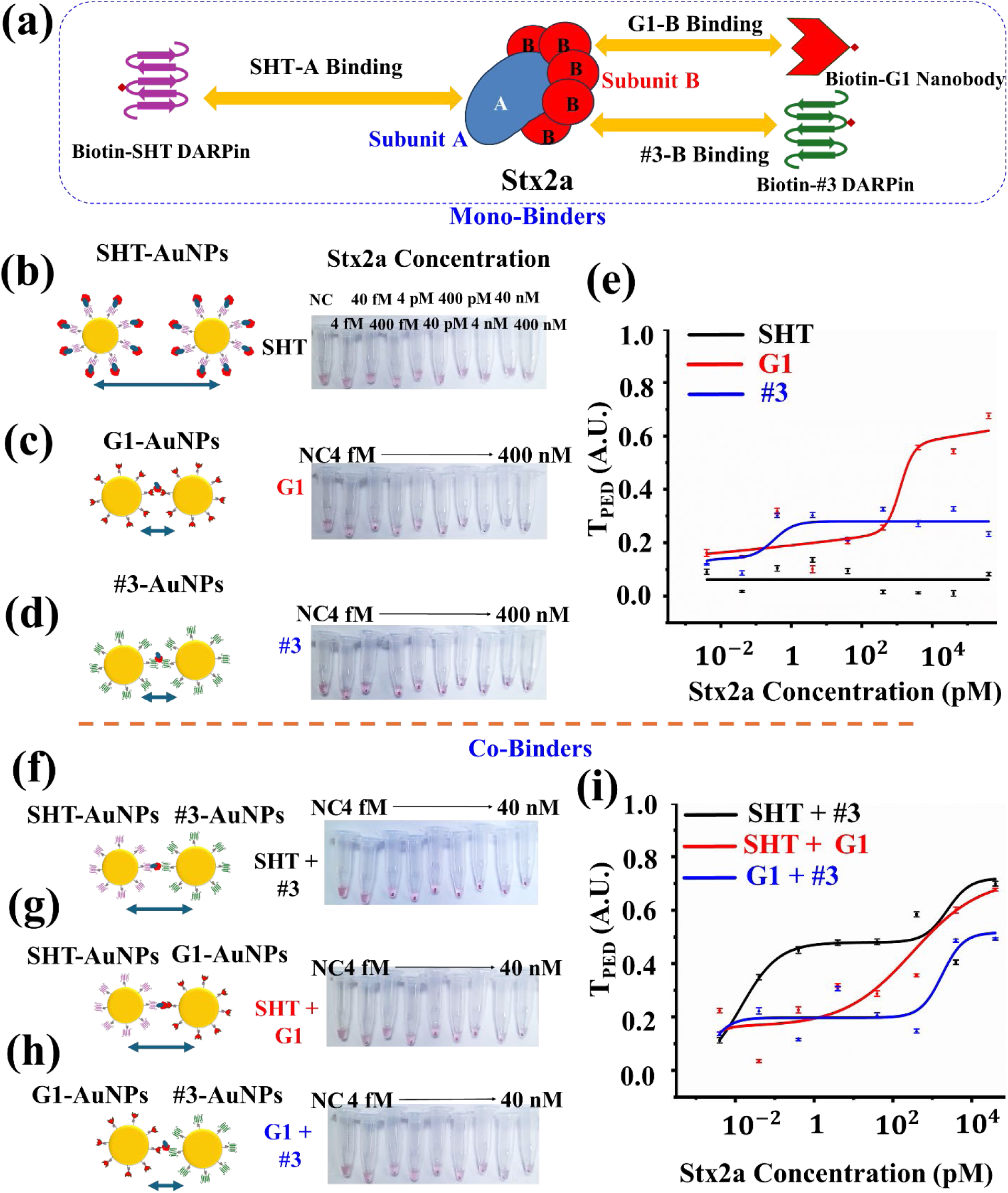
Co-binding impact on sensor performance. (a) Schematic representation of the Stx2a AB_5_ geometry and the binding SHT DARPin on A subunit and #3 DARPin and G1 nanobody binding to the B subunits of the toxin as rationale design for DARPin-RED platform binder selection. (b-d) Schematics of the hypothesized binding model and optical images of the testing tubes indicating Stx2a spiked in PBS samples ready for NasRED readout: Using mono-binders (b) SHT-AuNPs, (c) G1-AuNPs, and (d) #3-AuNps as sensor solution. NC indicates the negative control sample without Stx2 as control. (e) Extracted PED signals in PBS buffer using different mono-binder AuNP sensors, i.e. SHT-AuNPs (black solid line), G1-AuNPs (red solid line, LoD= 62 aM, or 6.2 fg/mL), #3-AuNPs (blue solid line, LoD=331 aM, or 33.1 fg/mL).. (f--i) Schematics of the hypothesized binding model and optical images of the testing tubes using co-binders: (f) SHT/#3-AuNPs, (g) SHT/G1-AuNPs, and (h) G1/#3-AuNps as sensor solution. NC indicates the negative control sample. (i) Extracted PED signals in PBS buffer using different co-binder AuNP sensors, i.e. SHT/#3-AuNPs (black solid line, LoD∼ 0.47 fM, or 47 fg/mL), SHT/G1-AuNPs (red solid line, LoD∼ 0.59 fM, or 59 fg/mL), G1/#3-AuNPs (blue solid line, LoD∼ 0.6 fM, or 60 fg/mL).

**Table 1.** Sensor performance analysis of Mono-binder and Co-binder Stx2a assays. mono-cycle protocol used for these assays.

| Performance metrics |  | Mono-binders |  |  | Co-binders |  |  |
| --- | --- | --- | --- | --- | --- | --- | --- |
|  |  | SHT | G1 | #3 | SHT/#3 | SHT/G1 | G1/#3 |
| Signal stability (RSS) |  | N/A* | 0.81 | 0.76 | 0.032 | 0.069 | 0.92 |
| Responsiveness<br>(Slopes) | r1 | N/A | 1.25 | 2.18 | 1.91 | 2.68 | 1.48 |
|  | r2 | N/A | 7.43 | 1.67 | 2.12 | 1.57 | 4.82 |
| Signal contrast (T<sub>Max</sub>) |  | N/A | 0.61 | 0.31 | 0.71 | 0.69 | 0.58 |
| Mean SNR |  | 7.82 | 37.89 | 32.79 | 45.14 | 47.01 | 43.14 |

\*N/A: SHT mono-binder yielded no significant signals.

We then assessed the sensor response to Stx2a-spiked PBSD samples using paired combinations of functionalized AuNPs (#3-AuNPs, G1-AuNPs, and SHT-AuNPs). Each reaction contained 9 µL each of functionalized AuNPs (18 µL total) and 6 µL of Stx2a-spiked PBSD, and the mixture was analyzed using the mono-cycle sensing protocol. SHT-containing co-binders exhibited superior sensor performance compared to mono-binders characterized by higher signal intensity (T_max_ ∼0.7 for SHT/#3 and SHT/G1 vs. T_max_ ∼0.6 for G1/G1 and G1/#3, and ∼0.3 for #3/#3), and reduced signal fluctuations (RSS ∼0.03 for SHT/#3 and ∼0.07 for SHT/G1 vs. RSS ∼0.8 for G1/G1 and #3/#3) (**Fig. 2 f-i**, **Table 1**, see methods). Additionally, the average signal to noise ratio (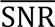) across tested concentrations exhibited an increased co-binder performance compared to mono-binders (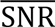 ∼45 for SHT/#3, ∼47 for SHT/G1 vs. 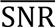∼37 for G1/G1 and ∼32 for #3/#3) (**Table 1**). This improvement is attributed to the cooperative binding of SHT (A-subunit) with either #3 or G1 (B-subunit) at spatially distant and non-overlapping sites, thereby mitigating steric hindrance and enhancing detection efficiency^42^. Conversely, the G1/#3 pair showed high signal variability (RSS∼0.9), likely due to competitive binding at adjacent B-subunit sites leading to steric hindrance and unstable cluster formation (**Fig. 2 h**, **Table 1)**. SHT/#3 combination was selected for further experiments due to its superior sensor performance (lowest RSS and high 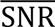). These results highlight the inherent trade-off between sensitivity and assay performance in molecular detection strategies.

### DARPin-RED performance in human samples and food matrices

To evaluate the potential of the DARPin-RED platform as POC and the impact of sample matrix on sensor performance, we applied this sensor for the detection of Stx2a in human (human pooled serum (HPS), whole blood) and food (lettuce juice extract, ground beef extract, and milk) samples. In these experiments, we first incubated Stx2a (1 µM) in the original human or food samples (as is) at room temperature for 30 minutes. Next, we prepared 4% of the human /food sample dilution in PBSD and used it in the serial dilution of pre-incubated Stx2a. Each sensing solution contained SHT-AuNP:#3-AuNP: diluted Stx2a at a 9:9:6 volume ratio in a total of 24 µL and was subjected to the mono-cycle sensing protocol.

DARPin-RED platform demonstrated high sensitivity (fg/mL) Stx2 detection in diverse human samples DARPin-RED. The LoDs of Stx2 calculated as 1.8 fg/mL (18 aM) in PBSD, 5.5 fg/mL (∼55 aM) in 4% HPS, and 427 fg/mL (∼4.3 fM) in 4% whole blood (**Fig. 3 a-d**), corresponding to ∼0.14, and 10.6 in 100% HPS, and whole blood, respectively. Matrix interferences were observed, reflected by variations in sensing performance indicators **(Table 2).** The LoD for Stx2a in blood-related samples (whole blood and serum) was reduced ∼3- and ∼25-fold compared to PBSD, likely due to the presence of human serum amyloid P component (HuSAP), an innate defense protein that binds, and partially neutralizes, Stx2^43^. The detection of Stx2a diluted in 20% whole blood or 100% HPS is possible, albeit with further reduced sensor performance than that diluted in 4% whole blood or HPS, likely due to the same HuSAP effect and/or optical interference from whole blood color (**Fig. S1**). The ability to detect Stx2a in 4% whole blood enables potential minimally invasive Stx2 detection using capillary blood sampling. The estimated Stx2a concentration in STEC patients is ∼2.6 pM in serum^44^ (assuming a molecular weight of ∼70 kDa). These concentrations are well above the LoDs attributed to undiluted matrices (1.3 fM in HPS, and 107.5 fM whole blood) by DARPin-RED, underscoring a great POC potential.

**Figure 3.**
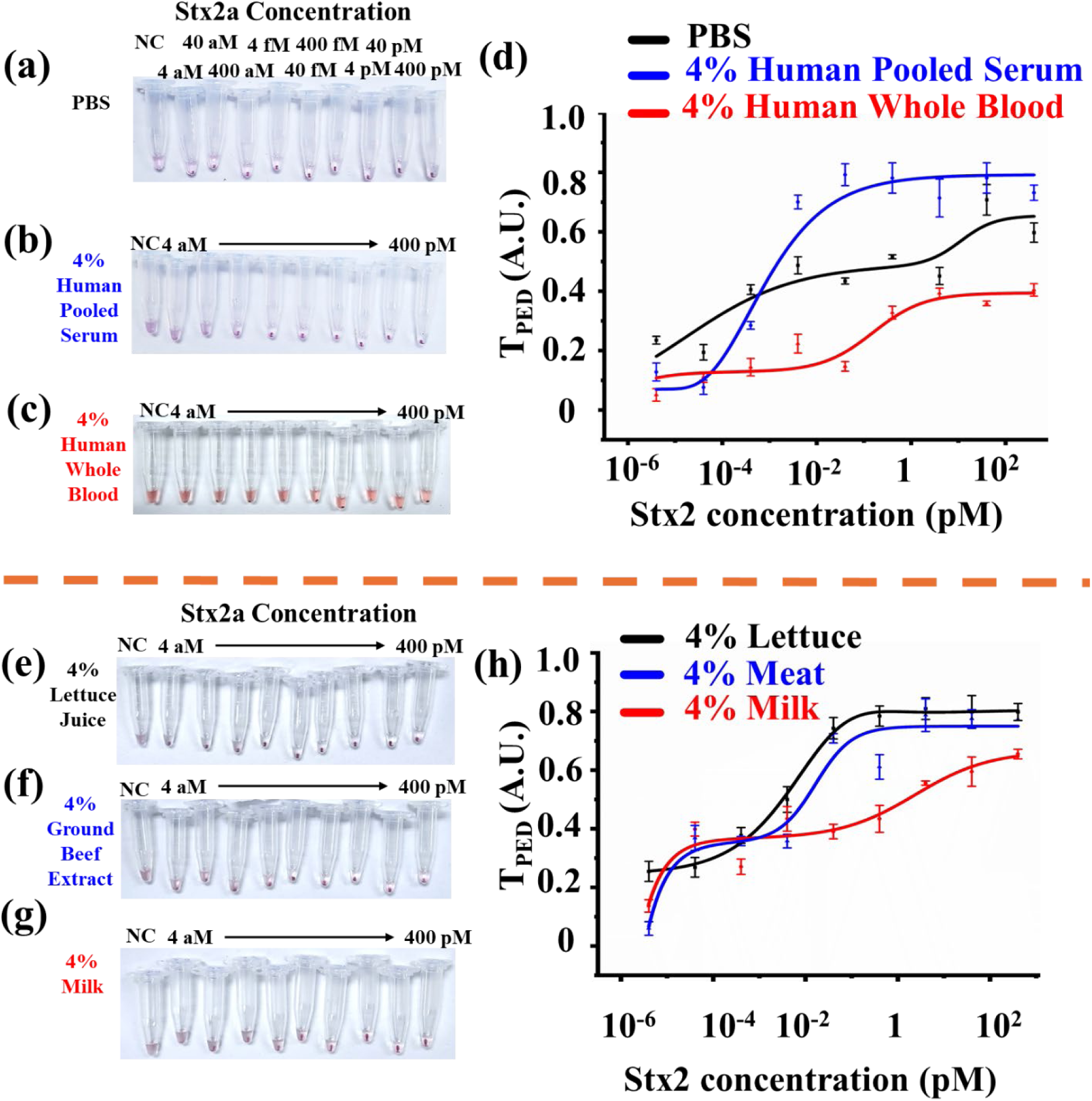
DARPin-RED performance in body fluids and food matrices. (a-c) Optical images of the testing tubes indicating Stx2a spiked in different human specimens ready for NasRED readout: (a) PBS, (b) human pooled serum, (c) human whole blood. All matrices diluted using PBS to reduce the matrix complexity. Stx2 concentrations on the tubes indicate the final concentration after dilution (see methods). NC indicates the negative control sample without Stx2 as control. (d) Extracted PED signals in different biological matrices, i.e. PBSD (black solid line, LoD∼ 18 aM, or 1.8 fg/mL), whole blood (red solid line, LoD∼ 4.27 fM, or 427fg/mL), HPS (blue solid line, LoD∼55 aM, or 5.5 fg/mL). (e-g) Optical images of the testing tubes indicating Stx2a spiked in different food matrices ready for NasRED readout: (e) lettuce juice extract, (f) beef, (g) milk. Food matrices diluted using PBS to reduce the matrix complexity. Stx2 concentrations on the tubes indicate the final concentration after dilution (see methods). NC indicates the negative control sample without Stx2 as control. (h) Extracted PED signals in different food matrices, i.e. lettuce juice extract (black solid line, LoD∼ 3 aM, or 0.2 fg/mL), milk (red solid line, LoD∼ 2 aM, or 0.2 fg/mL), and beef extract (blue solid line, LoD∼ 4.7 aM, or 0.47 fg/mL).

**Table 2.** Sensor performance analysis of biological and food matrices impact under mono-cycle protocol. All matrices analyzed at 4%.

| Performance metrics |  | Biological matrices |  |  |  | Food matrices |  |  |  |  |
| --- | --- | --- | --- | --- | --- | --- | --- | --- | --- | --- |
|  |  | PBS | HPS | Whole blood | Mono-Cycle |  |  | Dual-Cycle |  |  |
|  |  |  |  |  | Lettuce | Milk | Beef | Lettuce | Milk | Beef |
| Sensitivity (LoD, aM) |  | 18 | 55 | 4270 | 3 | 2 | 4.7 | 600 | 16200 | 1620 |
| Signal stability (RSS) |  | 0.023 | 0.031 | 0.050 | 0.032 | 0.072 | 0.034 | 0.012 | 0.014 | 0.014 |
| Responsiveness<br>(Slopes) | r1 | 0.4 | 0.9 | 0.8 | 2.5 | 1.4 | 2.4 | 0.56 | 0.47 | 0.54 |
|  | r2 | 1.2 | 0.5 | 0.8 | 0.52 | 0.31 | 0.43 | 3.5 | 2.8 | 3.5 |
| Signal contrast (T<sub>Max</sub>) |  | 0.65 | 0.79 | 0.39 | 0.83 | 0.68 | 0.79 | 0.77 | 0.64 | 0.73 |
| Mean SNR |  | 26.36 | 16.83 | 14.84 | 16.42 | 18.49 | 21.95 | 8.45 | 17.65 | 10.79 |

To evaluate the potential application of DARPin-RED for food safety surveillance programs, we assessed the interference of food matrices. Filtered extracts of lettuce, ground beef, and milk were spiked with 1 µM Stx2a and incubated at room temperature for 30 minutes before being serially diluted in PBSD supplemented with 4% food matrix. For each sensing experiment, 6 µL diluted Stx2a was mixed with 9 µL each of SHT/#3-AuNPs (total 24 µL), and the mixture was subjected to dual-cycle sensing protocol to improve the signal resolution. Highly sensitive detection of Stx2 was observed in 4% lettuce and ground beef extracts, as well as 4% milk, reaching LoDs of 0.3, 0.47, and 0.2 fg/ml, respectively, which corresponds to 7.5, 11.25, and 5 fg/ml in undiluted lettuce, beef extract, and milk. A slightly higher noise (RSS ∼ 0.07) and decrease in signal intensity (T_max_∼0.68) was observed in diluted milk compared to lettuce/beef extract (RSS ∼0.03, T_max_ ∼0.8), possibly due to protein interference and/or milk composition elements^45^ **(Fig. 3 e-h**, **Table 2)**.

Overall, the DARPin-RED platform demonstrated highly sensitive (aM to fM LoDs) for Stx2 in diverse clinical and food samples while maintaining assay robustness (**Table 2**). This capability underscores DARPin-RED versatility in rapid toxin detection with minimal sample preparation.

### DARPin-RED performance against different toxin subtypes

Fifteen subtypes of Stx2 have been reported (a-o) to date^46^, with subtypes Stx2a, Stx2c and Stx2d most commonly associated with severe human illness and subtypes Stx2e, Stx2f and Stx2g primarily associated with animal diseases^46^. The amino acid identity between different subtypes range between 69.7-92.9% for the A subunit and 67.2-91.3% for the B subunit^46^. Although DARPin SHT and #3 were engineered using Stx2a as the target, due to the relatively high sequence identity, they potentially also bind other Stx2 subtypes, prompting us to evaluate the ability of DARPin-RED to detect different Stx2 subtypes a-g.

To account for potential interference from human HuSAP in toxin subtype detection, we detected the different Stx2 subtypes in both PBSD and PBSD supplemented with 4% HPS. To ensure comparability of signal intensity across different subtypes, a standard control (SC) was established using recombinantly purified Stx2a at 10 ng/mL. Different Stx2 subtypes were diluted to 10 ng/mL in PBSD or PBSD supplemented with 4% HPS. All tested Stx2 subtypes showed higher T_PED_ signal than the negative control (NC), suggesting their detection by DARPin-RED (**Fig. 4 a,b**). The presence of HuSAP did not impact the detection except for Stx2g, whose detection signal was significantly reduced in 4% serum than in PBSD. The T_PED_ signal for Stx2b is approximately half of SC (T_max_ ∼ 0.2 for Stx2b vs. 0.4 for SC), while Stx2f was almost undetectable (T_max_ ∼ 0.05). The variations in signal intensity observed among Stx2 subtypes can be attributed to specific amino acid differences between them^47^. These structural variations can affect the toxin’s interaction with host receptors and its immunogenic properties, as well as their affinity to SHT/#3.

**Figure 4.**
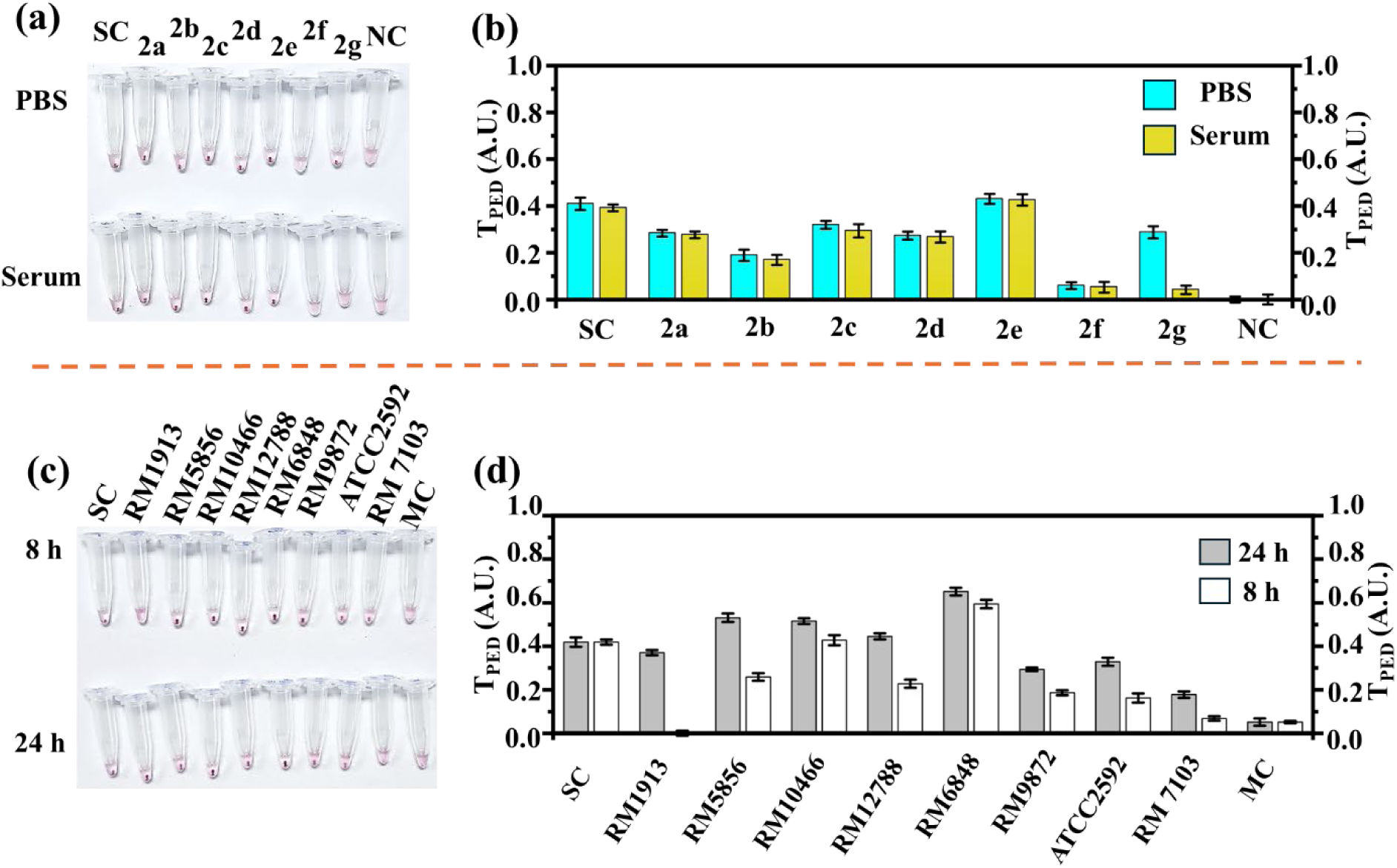
Detecting toxin subtypes and bacterial strains. (a) Optical images of the testing tubes indicating Stx2 subtypes (a-g) spiked PBS and serum ready for NasRED readout. NC indicates the negative control sample without Stx2 as control. SC indicates the subtype control containing Stx2a by which the DARPins were developed. All samples spiked at the concentration of 10 ng/mL. (b) Extracted PED signals in PBS and serum matrices plotted against the subtypes (a-g) and negative and standard controls (NC and SC). (c) Optical images of the testing tubes indicating STEC variants cultured sample after 8 and 24 hours of culturing ready for NasRED readout. MC indicates the matrix control of culturing medium without Stx2 as control. CC indicates the concentration control containing Stx2a by which the DARPins were developed at 10 ng/mL. (d) Extracted PED signals bacterial culture matrix plotted against the STEC variants and matrix and standard controls (MC and SC).

Different STEC variants produce different Stx2 subtypes at varying levels, and some also co-express Stx1 (e.g., RM12788, **Table 3**). We further assessed the sensor’s performance in detecting different STEC strains in culture media. To validate the authenticity of sensor signals in regard to matrix interferences and subtype signal variations, we included a matrix control (MC) comprising of culture medium alone and a standard control (SC) consisting of 10 ng/mL of Stx2a. Bacterial cultures were harvested at 8- and 24-hours post-inoculation and filtered to remove bacterial debris (see methods). For biosensor testing, 6 µL culture supernatant was mixed with 9 µL each of SHT-/#3-AuNPs (total 24 µL), and the mixture was subjected to mono-cycle sensing protocol. Our DARPin-RED platform successfully detected toxins in all the tested cultures (**Fig. 4 c,d**). In some cases, such as RM1913, RM12788, and RM5856, the 24-hour culture yielded a much higher T_PED_ signal than the 8-hour cultures, likely indicating the accumulation of secreted toxin in these cultures over time. Negligible signals were observed for ATCC 25922 and RM7103 (Stx-negative controls). These results highlight the platform’s potential for sensitive early detection of different STEC infections in 8 hours of culturing.

**Table 3.**
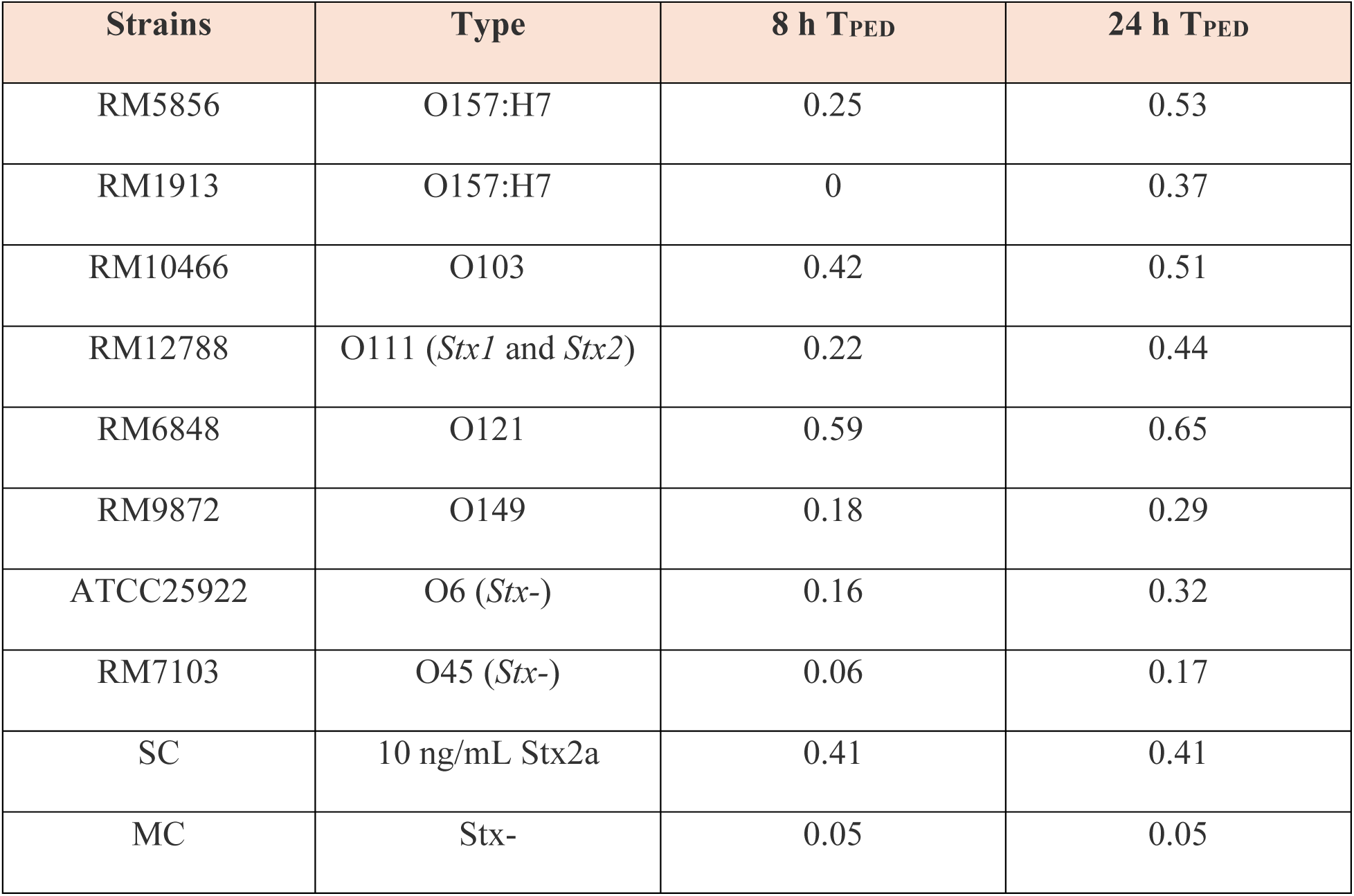
Detection of Stx2 in culture supernatants at 8- and 24-h post-inoculation.

| Strains | Type | 8 h T<sub>PED</sub> | 24 h T<sub>PED</sub> |
| --- | --- | --- | --- |
| RM5856 | O157:H7 | 0.25 | 0.53 |
| RM1913 | O157:H7 | 0 | 0.37 |
| RM10466 | O103 | 0.42 | 0.51 |
| RM12788 | O111 (*Stx1* and *Stx2*) | 0.22 | 0.44 |
| RM6848 | O121 | 0.59 | 0.65 |
| RM9872 | O149 | 0.18 | 0.29 |
| ATCC25922 | O6 (*Stx*-) | 0.16 | 0.32 |
| RM7103 | O45 (*Stx*-) | 0.06 | 0.17 |
| SC | 10 ng/mL Stx2a | 0.41 | 0.41 |
| MC | Stx- | 0.05 | 0.05 |

### Detection of Stx2 in Cultured Food Samples Using the Mono-Cycle and Dual-Cycle NasRED Protocols

To evaluate the sensor’s capability to identify STEC contamination in complex food samples based on the production of Stx2, STEC were spiked in lettuce, milk, and ground beef (0.4 CFU/mL or 0.4 CFU/g, **Table 4**). 25 mL/g of the bacteria-spiked food samples was mixed with 74 mL of growth media (modified tryptone soy broth with 100 ng/mL mitomycin C) and cultured at 42 °C with shaking (200 rpm) for 8 and 16 hours. The liquid culture supernatant was harvested by centrifugation, serially diluted in PBSD supplemented with 4% of the respective food samples and analyzed using both mono-cycle and dual-cycle NasRED protocols. Samples containing serially diluted Stx2a in PBSD supplemented with 4% of respective food samples were used to generate the calibration curves for each protocol (**Fig 5 a-d**). The Stx2a concentration in STEC-inoculated samples was quantified by mapping sensor signal intensities to the corresponding calibration curves (**Fig 5 e, f**). The baseline was determined as the highest quantified value obtained from the matrix control (BPW) and Stx2-negative strain (ATCC 25922), accounting for the matrix interference and quantification error. Statistical analysis was performed using Tukey’s two-way ANOVA for paired comparisons of experimental factors, assessing the sensor’s ability to distinguish Stx2STEC from non-STEC strains and the impact of bacterial incubation time and inoculation dose on toxin detection (see methods).

**Figure 5.**
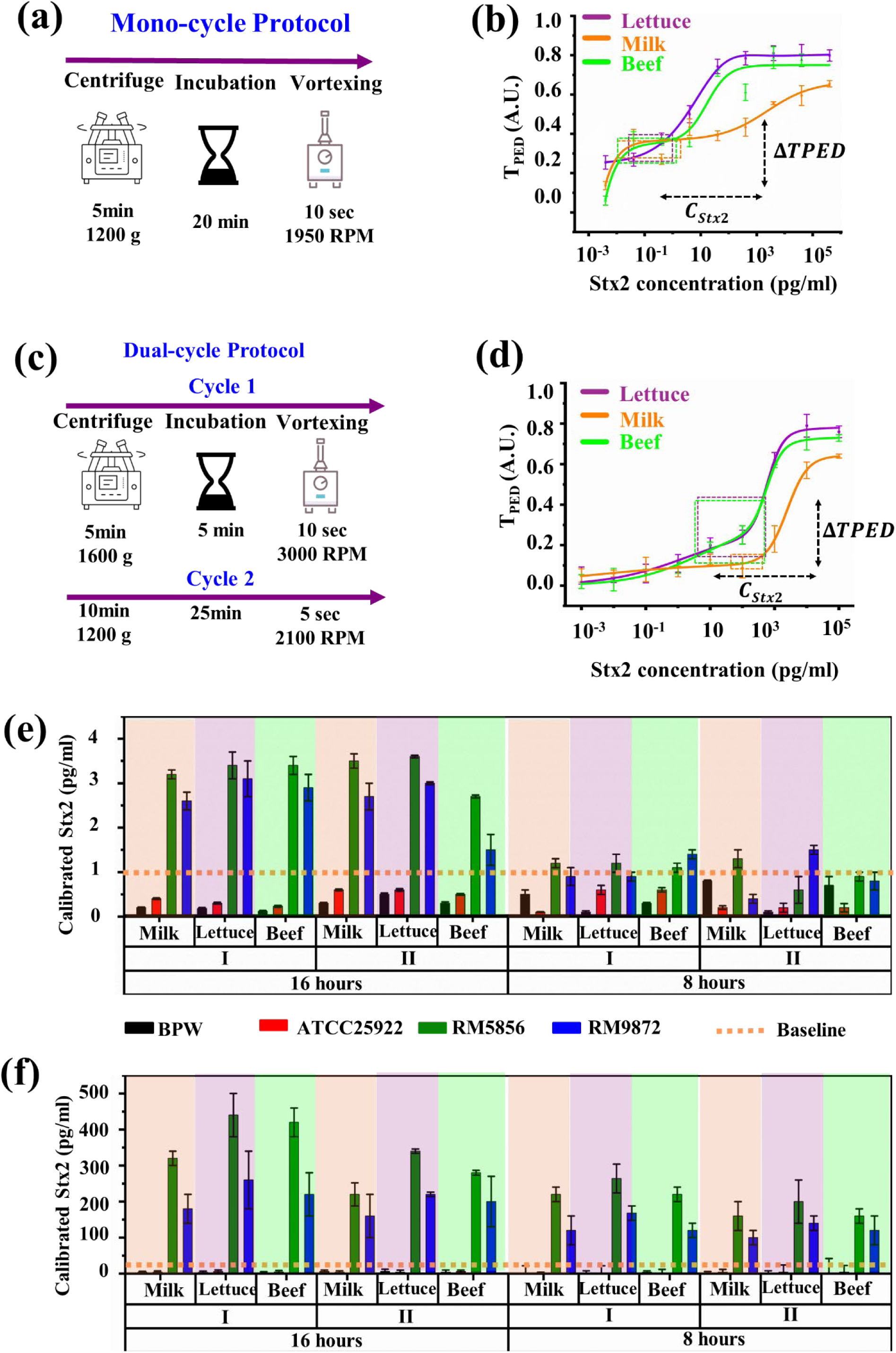
Evaluation of Cultured Food Sample Detection Using Different Protocols. (a) Schematic representation of the mono-cycle protocol. (b) Extracted sensor response curves for food samples spiked at 4% matrix concentration, analyzed using the mono-cycle protocol in lettuce juice, milk, and ground beef extract. The quantified concentration for inoculated samples (C_Stx2_) and their corresponding signals visualized as dashed boxes on the calibration curves. (c) Schematic representation of the dual-cycle NaSRED protocol. (d) Extracted sensor response curves for food samples spiked at 4% matrix concentration, analyzed using the dual-cycle protocol in lettuce juice, milk, and ground beef extract. The quantified concentration for inoculated samples (C_Stx2_) and their corresponding signals (ΔT_PED_) visualized as dashed boxes on the calibration curves. _(_(e-f) Calibrated toxin concentrations in food samples plotted against matrix type, inoculation state, and incubation time for (e) mono cycle protocol and (f) dual cycle protocol. The baselines (1 fg/mL for mono-cycle protocol and 6 pg/mL for dual-cycle protocol) represent the highest sensor noise level, determined by the Stx2-negative variant (E. coli ATCC 25922) and matrix control (BPW).

**Table 4.** Actual inoculum of E. coli strains used to spike food matrices for enrichment with mitomycin C for 8– or –16 h and calculated from serially diluted agar plate counts.

| Strain | Serotype | Stx2 Expression | Experiment I<br>(CFU/g/mL) | Experiment II<br>(CFU/g/mL) |
| --- | --- | --- | --- | --- |
| ATCC25922 | O6, stx- | None | 0.23 | 0.19 |
| RM5856 | O121:H19 | High | 0.38 | 0.19 |
| RM9872 | O145 | Low | 0.19 | 0.14 |

In the mono-cycle protocol, extracted signals showed a clear distinction between Stx2-positive and Stx2-negative strains, with RM5856 (high Stx2 producer) and RM9872 (low Stx2 producer) exhibiting higher sensor signals compared to the non-toxigenic ATCC 25922 strain and matrix control (BPW) (**Fig. 6 b**, p< 0.001). Also, for RM5856 and RM9872, a higher concentration of Stx2 is detected at 16 hours than 8-hours post inoculation (**Fig. S25 b**, p> 0.05). However, the protocol failed to differentiate between high- and low-yield producers, as signal intensities for RM5856 (high yield) and RM9872 (low yield) were not significantly different (**Fig. 6 b**, p > 0.05) and negative control samples were not significantly different from the baseline (1 pg/mL, **Fig. 5 e**). Furthermore, the food matrix had no significant effect on sensor response (**Fig. S2 a**, p > 0.05), supporting the robustness of the platform across different food matrices. However, the mono-cycle protocol could not differentiate between different inoculation concentrations (**Fig. 6 a**, p > 0.05), limiting its potential for precise bacterial load estimation, which may be critical for predicting the virulence of the RM5856. We noticed that the mono-cycle calibration curves followed the biphasic dose response model in which the sensor yields signals at low concentrations (fg/mL range) but the middle range concentrations (pg/mL range) fall in the plateau of the curve (**Fig. 5 b**). Deducing from our previous work^37^, we hypothesized that it is due to the increase in the number of clusters rather than the size of each cluster at these middle-range concentrations. These smaller clusters are more susceptible to resuspension by vortex agitation than larger clusters, leading to weak T_PED_ signal changes at middle-range concentrations.

**Figure 6.**
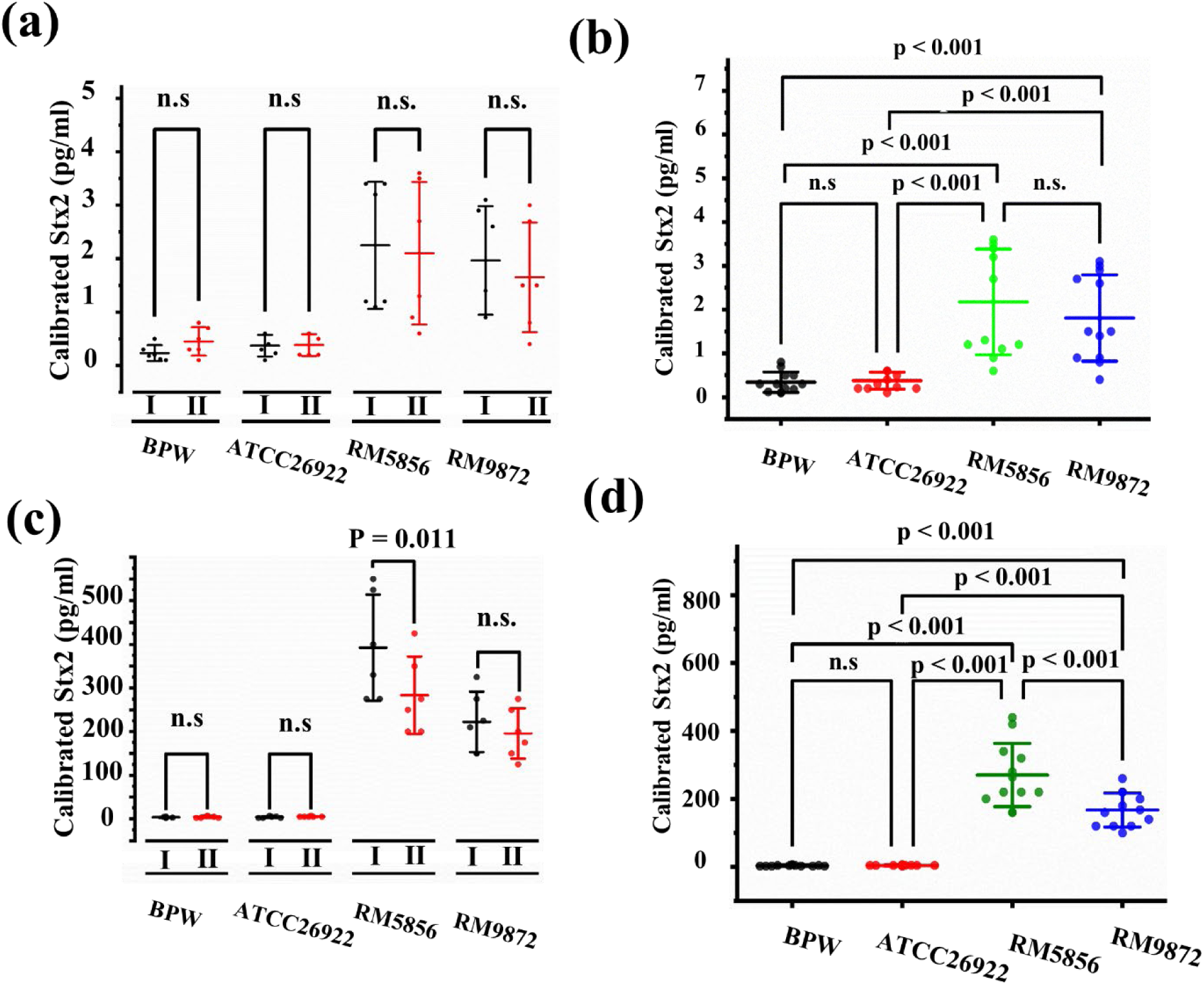
Statistical Analysis of the STEC Inoculated Food Samples Quantification Performance. (a) ANOVA analysis of inoculation concentrations and bacterial strains, indicating that the mono-cycle protocol does not differentiate bacterial inoculation states. (b) Comparative analysis of bacterial strains, showing the sensor’s ability to distinguish Stx2-producing from non-producing strains but not between high-yield (RM 5856) and low-yield (RM 9872) Stx2 producers. (c) ANOVA analysis of bacterial strains and inoculation conditions, demonstrating that the dual-cycle protocol differentiates bacterial inoculation doses for high-yield Stx2 producers (RM 5856, p < 0.011). (d) Further analysis of bacterial strains, showing that the dual-cycle protocol distinguishes high-yield (RM 5856) from low-yield (RM 9872) Stx2 producers (p < 0.001).

We developed a dual-cycle sensing protocol to enhance the sensor’s ability to distinguish different middle-range Stx2a concentrations. Unlike the mono-cycle sensing protocol, which consists of a single round of centrifugation, incubation, and vortexing (**Fig. 5a**), the dual-cycle sensing protocol contains two rounds of centrifugation, incubation, and vortexing with AuNPs oligo-clusters formed during the first round act as seeds in the 2^nd^ round, facilitating further interactions with remaining targets in solution (**Fig. 5 c**). The dual-cycle sensing protocol achieved improved curve smoothness at concentrations between pg/mL and ng/mL (r ∼ 0.5, **Fig. 5 d**) despite a slight loss of sensitivity (> pg/mL LoDs). This protocol also significantly enhanced signal resolution, yielding a significantly lower baseline (6 pg/mL) compared to Stx2-producing samples (>200 pg/mL, **Fig. 5 f**). Statistical analysis confirmed a significant difference (p < 0.001) in sensor response between high-yield (RM5856) and low-yield (RM9872) Stx2-producing strains (**Fig. 6 d**), a distinction not observed in the mono-cycle protocol. Additionally, the dual-cycle approach enabled the differentiation of bacterial inoculation doses for high-yield Stx2 producers (RM5856, p = 0.011, **Fig. 6 c**), demonstrating its potential for semi-quantitative toxin detection. A key advantage of the dual-cycle sensing protocol is its ability to differentiate Stx2 production within 8 hours post STEC inoculation from negative controls (**Fig. 5 f**), thereby significantly reducing the turnaround time compared to conventional culturing methods requiring over 24 hours^33^. The significant differences in sensor response between 8-hour and 16-hour incubation times (p < 0.001, **Fig. S2 d**) underscores its potential for early-stage pathogen detection. Notably, Stx2-negative strains (ATCC 25922) and the matrix control (BPW) produced the lowest signals **(Fig. 5 f)**, reinforcing assay specificity.

Overall, these findings demonstrate that the dual-cycle NasRED protocol provides superior resolution, specificity, and differentiation between Stx2-producing and non-producing strains compared to the mono-cycle approach. While the mono-cycle protocol was effective for qualitative detection, its inability to distinguish between strains with different toxin-production yields and cultures inoculated with different bacteria doses, and low signal resolution at 8-hour samples limit its quantitative applications. The dual-cycle protocol overcomes these limitations, making it a more suitable approach for early toxin detection in foodborne pathogen surveillance.

## Conclusion

This study presents DARPin-RED, a high-sensitivity POC diagnostic platform for Stx2 detection in clinical and food safety applications. By integrating nanoparticle-assisted detection with DARPins, the system enables rapid, label-free, and wash-free toxin quantification with attomolar sensitivity. The dual-cycle NasRED protocol, developed to address quantification resolution mid-range concentrations, significantly improved its capability of differentiating high toxin producer (RM5856) and low toxin-producer (RM9872) STEC strains (p < 0.001) and enhanced its dose-response function (p = 0.011), confirming its quantitative diagnostic capabilities. The platform demonstrated its robusticity for the detection of Stx2 across multiple samples, including clinical samples such as serum, and whole blood with LoDs of 137, and 84 fg/mL, respectively, and complex food samples such as lettuce, milk, and meat with LoDs of 7.5, 5, 12 fg/mL, respectively (Table 5). More importantly, the sensor developed in this study minimized the HuSAP interference, which was a critical issue in other detection platforms. The system’s ability to distinguish Stx2-positive from Stx2-negative strains with high precision, even in the presence of food matrix effects, underscores its potential for real-world deployment. A key advancement of DARPin-RED is its ability to detect Stx22 within 8 hours after bacterial incubation, a major improvement over conventional methods, which require >24 hours for toxin detection^28^. This high sensitivity is critical for early intervention, particularly in clinical settings where timely diagnosis can enable prompt medical decision-making and reduce the risk of HUS progression. The platform’s high sensitivity also enables direct toxin detection in biological specimens, bypassing the need for bacterial enrichment, which remains a bottleneck in standard STEC diagnostics^48^. In addition to its clinical implications, DARPin-RED provides a scalable and cost-effective solution for STEC surveillance in food safety applications. The ability to rapidly screen high-risk food products with minimal sample preparation is essential for product recall and preventing STEC outbreaks and ensuring compliance with food safety regulations. Furthermore, its portability and ease of use make it well-suited for on-site testing in decentralized settings, reducing dependence on centralized laboratories and accelerating outbreak^49^.

**Table 5.**
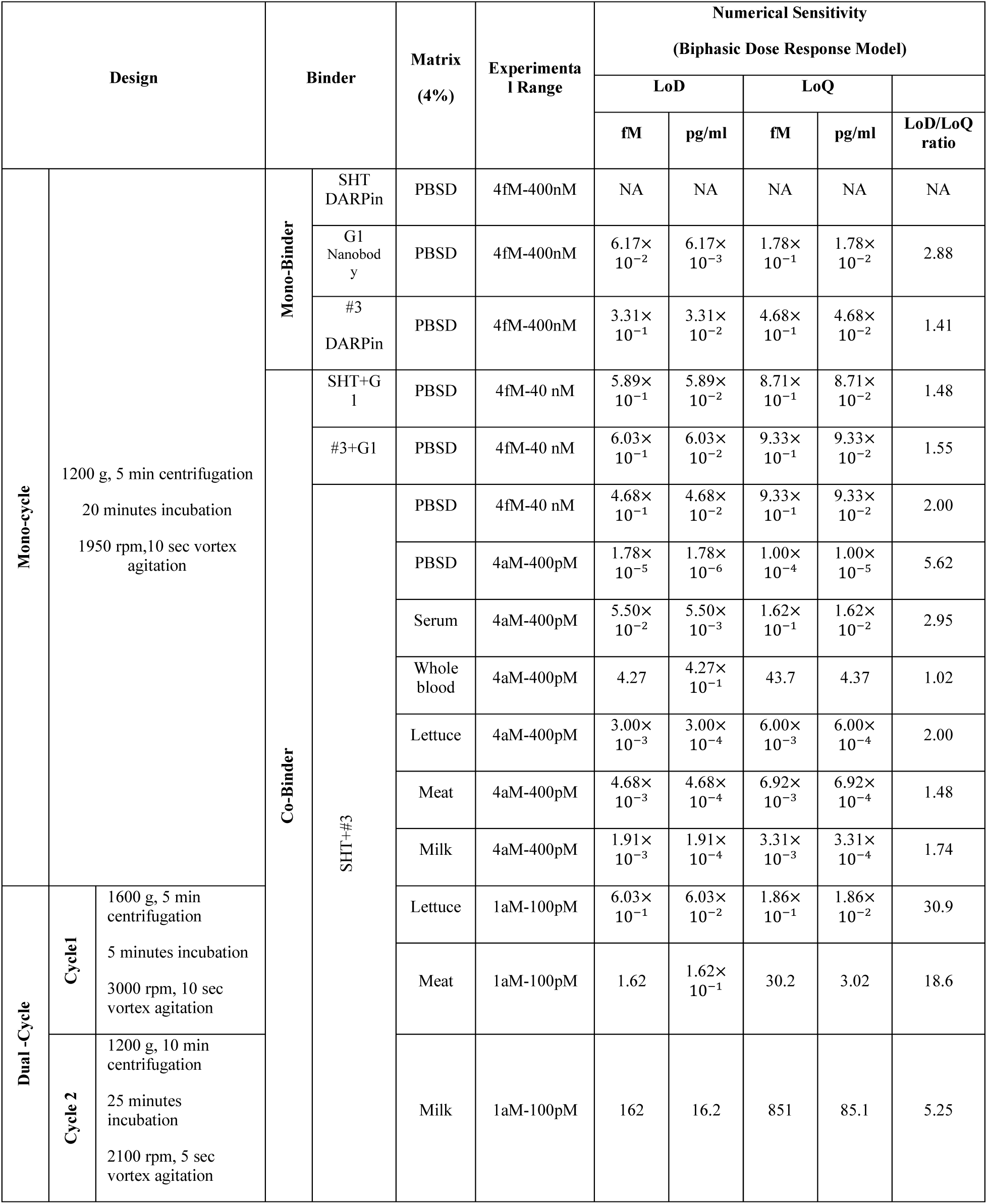
DARPin-RED design and numerical sensitivity analysis in detecting Stx2.

## Methodology

### Instrumentation Sources

The microcentrifuge (accuSpin Micro 17) and vortex mixer Digital Vortex Mixer (Catalog No. 02-215-418) were sourced from Thermo Fisher.

### Stx2a Standard and DARPin Preparation

#### Stx2a Standard Preparation

The Stx2a standard and DARPins used in this study were obtained through recombinant expression and purification, following established protocols in our previous study^31^.

Recombinant Stx2a holotoxin, was expressed in E. coli BL21(DE3) cells and purified using nickel-nitrilotriacetic acid (Ni-NTA) affinity chromatography. The purified Stx2a holotoxin was concentrated and buffer-exchanged into 1× PBS (pH 7.4) using an Amicon ultrafiltration unit (MWCO 50 kDa) to remove excess salts and ensure compatibility with the assay conditions. Protein purity was verified by SDS-PAGE analysis, and the final protein concentration was determined using the Bicinchoninic Acid (BCA) assay (Thermo Scientific). Purified Stx2a was stored at −20°C in 50% glycerol until use.

### DARPin Expression and Purification

The DARPins (DSHT and D#3) used for biosensor functionalization were selected from an in-house DARPin library (∼2 × 10⁹ variants) developed through phage panning and directed evolution^50,51^. The phage panning process involved biotinylation of Stx2a using EZ-Link-Sulfo NHS-LC biotin (Pierce), followed by incubation with the phage-displayed DARPin library. High-affinity binders were selected based on binding specificity, stability, and off-rate kinetics.

Candidate DARPin clones were expressed in E. coli BL21(DE3) cells, purified via immobilized metal affinity chromatography (IMAC) using a His-tag purification system, and subjected to endotoxin removal using Pierce™ High-Capacity Endotoxin Removal Spin Columns (Thermo Scientific). Endotoxin levels in the final purified proteins were quantified using the Pierce™ Chromogenic Endotoxin Quant Kit (Thermo Scientific) and maintained at ≤14 EU/mL to ensure compatibility with biosensor assays. Purified biotinylated DARPins were stored in DPBS (Fisher Scientific) at −80°C and used for functionalization of streptavidin-coated AuNPs in the biosensing experiments.

### Electronic Readout System

The PED system, similar to our recent study^37^, comprised of Light-emitting diode (LED) (WP7113PGD, Kingbright), photodiode-integrated circuit (SEN-12787, SparkFun Electronics), incorporating a digital light sensor (APDS-9960, Broadcom), custom-fabricated tube holder, produced using a 3D printer (Qidi Tech X-Plus) with black carbon fiber polycarbonate filament. The tube holder was specifically designed to securely accommodate standard 0.5 mL Labcon super clear microcentrifuge tubes (Labcon inc. cat# 3046-870-000-9), ensuring precise optical alignment. The photodiode APDS-9960 was operated with a bias voltage of 3.3 V and interfaced with a microcontroller (Atmega328) for signal processing and digitization. This setup provided reliable electronic readouts of light intensity variations with minimal noise. All electronic components, unless otherwise specified, were procured from DigiKey, and their integration was optimized to ensure consistent performance in the detection system.

### 1× PBS Dilution (PBSD) Buffer Preparation

1× PBS dilution buffer was prepared by mixing 10× phosphate-buffered saline (PBS) (Fisher Scientific) with glycerol (Sigma-Aldrich), bovine serum albumin (BSA) (Sigma-Aldrich), and deionized water (Fisher Scientific). The final composition of the dilution buffer was 1× PBS, 20% (v/v) glycerol, and 1 wt% BSA, with a pH of approximately 7.4. This buffer was used for the preparation of AuNP sensors, toxin serial dilutions, and diluted biological and food media to ensure stability and consistency in the experimental setup.

### Sensing Solution Preparation and Quantification

To functionalize streptavidin gold nanoparticles (AuNPs) (∼0.13 nM, 80 nm, OD10, CAT#: ACC-80-04-15, Cytodiagnostics Inc.) with DARPin, 50 μL of streptavidin-coated AuNPs (OD10) was incubated with 20 μL of biotinylated DARPins (∼1.2 μM) at room temperature for 2 hours to allow for biotin-streptavidin binding. Next, 1000 μL of PBSD was added to the mixture and the mixture was centrifuged at 9,600 g for 10 minutes to pellet the functionalized AuNPs. Supernatant (1050 μL) was removed and replaced with an equal volume (1050 μL) of PBSD. This washing step was repeated twice to ensure the total removal of unbound biotinylated DARPin. The purified sensing solutions were measured using PED to determine the AuNP ^OD^PED, and then adjusted to the desired concentration mixing with PBSD (e.g., ^OD^PED∼ 0.5, ∼0.019 nM for 80 nm AuNPs). Prior to each sensing experiment, the stock sensing solutions were aliquoted into 1.5 mL Eppendorf tubes (500 µL/tube) and stored at 4C for 3 days to ensure stability and consistency.

### ^OD^PED quantification

For background normalization, the transmission signals of 24 μL PBSD in 0.5 mL microcentrifuge tubes were recorded using an in-house PED device, and the average transmission value, was determined by calculating the mean of five measurements taken along five random orientations 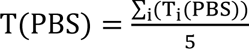 (𝑖=1 to 5). Similarly, the transmission values of the functionalized AuNP solution were measured under identical conditions, averaged across the same five orientations, and recorded as 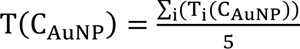 (i=1 to 5) at the corresponding AuNP concentration (C_AuNP_). The PED optical density, was then computed based on PED signal output using:

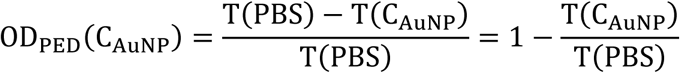

### Stx2 Detection in Spiked Samples

#### DARPin-RED Sensing Experiment

A 9 μL aliquot of each functionalized DARPin-AuNPs (total of 18 μL, OD_PED_∼0.5 AuNPs, in PBSD) was mixed with 6 μL of target sample to achieve a final 24 μL reaction volume (9:9:6 DARPin-AuNP to target ratio leading to 4-fold dilution of the target sample). The mixture was vortex agitated at 2000 RPM for 5 seconds to facilitate interaction and processed according to mono- or dual-sensing protocols. For Stx2a standard curve development, recombinant Stx2a was serially diluted from 400 nM to 4 aM in PBSD. The final concentration of Stx2a in the sensing solution ranges between 100 nM to 1 aM.

### Biological Media Preparation

Human pooled serum (HPS) and single-donor whole blood (WB) were obtained from Innovative Research, Inc. and used at a final concentration of 4%, diluted with PBSD. To evaluate the sensitivity of DARPin-RED for HPS, and blood samples, recombinant Stx2s (final 1 µM) was first incubated in the 100% (as is) serum, and blood samples, at room temperature for 30 minutes, then diluted with 4% of HPS or blood samples to achieve the desired final target concentrations and used in DARPin-RED sensing experiment following the 9:9:6 DARPin-AuNP to target ratio.

### Food Matrix Preparation

Food matrices, including lettuce juice, ground beef extract, and milk, were processed to ensure compatibility with the sensing assay while minimizing matrix interference. Lettuce juice extract was prepared by crushing 5 g of lettuce leaves in a mortar, followed by centrifugation at 12,000 g for 5 minutes. The supernatant was collected and stored at −80 °C until use. Ground beef extract was prepared from 70% lean ground beef. 40 mg of beef was resuspended in 1 mL of PBSD. The mixture was centrifuged at 12,000 g for 5 minutes, and the supernatant was collected and stored at −80 °C until use. Milk samples were prepared by diluting whole milk (5% fat) with to 4% using PBSD. 4% food matrix was prepared by diluting each food matrix 25-fold in PBSD. To evaluate the sensitivity of DARPin-RED in diluted food matrices, recombinant Stx2s (final 1 µM) was first incubated with the respective food matrix (as is) at room temperature for 30 minutes, then serially diluted in 4% food matrix and used in the sensing experiments following the same 9:9:6 DARPin-AuNP to target ratio.

### Detection of Stx2 in STEC Supernatants

Bacterial strains were streaked from glycerol stocks onto Luria Broth (LB) agar plates under biosafety conditions and incubated at 37 °C overnight^47^. The strains used in this study included Stx2-producing (RM5856 (O121, high Stx2 producer), RM9872 (O145, low Stx2 producer), RM1913, RM10466, RM12788, RM6848 and RM7013 and Stx2-negative *Escherichia coli* (ATCC25922 (O6, STX-negative control)) (Table 3). Single colonies from each plate were inoculated into 15 mL Falcon tubes containing 10 mL LB supplemented with 100 ng/mL mitomycin C and incubated at 37 °C with shaking at 200 rpm. At 8- and 24-hours post-inoculation, 4 mL of each bacterial culture were collected and centrifuged at 10,000× g for 10 minutes at 4 °C to pellet bacterial cells. The supernatants were filtered through a 0.2 μm membrane filter to remove the residual cells, aliquoted and stored at −80 °C until use. For biosensor testing, undiluted supernatant was used following the 9:9:6 DARPin-AuNP to target ratio, as previously described.

### Detection of Natively Expressed Stx2 in STEC-Spiked Food

Single colonies of Stx2-positive (RM5856 (O121:H19, high Stx2 producer), and RM9872 (O145, low Stx2 producer)) and Stx-negative *E. coli* strains (ATCC25922 (O6, Stx-negative)) were inoculated into tryptone soy broth (TSB, Fisher Scientific) and incubated at 37 °C for 18 hours^45^. Overnight cultures were then serially diluted to achieve a desired 10 colony-forming unit (CFU) per 25 g (or mL) of food sample. The actual inoculum levels were later determined by serially diluting each sample in TSB (Fisher Scientific) and plating 0.1 mL of each dilution onto Tryptic Soy Agar plates (TSA, Fisher Scientific) for manual colony counting after overnight incubation at 37 °C. Each food sample (25 g or 25 mL) was spiked with 1 mL of diluted bacteria (10 CFU/mL) or a buffered peptone water (BPW, Fisher Scientific) and homogenized in stomacher bags (∼0.4 CFU/g or 0.4 CFU/mL final concentration). For enrichment experiments, 74 mL of modified tryptone soy broth (mTSB, Neogen) containing 100 ng/mL mitomycin C (Fisher Scientific) was added to each sample (25 g or 25 mL) immediately before homogenization. The sample-broth mixture was homogenized using a stomacher paddle blender for 2 minutes at 230 rpm and incubated at 42 °C with shaking at 200 rpm for 8 and 16 hours to allow bacterial growth and Stx2 production. Post-incubation, 2 mL of liquid culture from each stomacher bags was transferred to microcentrifuge tubes (USA Scientific) and centrifuged at 10,000× g for 10 minutes at 4 °C. The supernatants were collected and filtered through a 0.2 µm low-binding membrane filter (Millex, San Jose, CA, USA) to remove residual bacterial cells. The cleared supernatants were aliquoted into 1.5 mL tubes and stored at −80 °C until analysis using the DARPin-RED biosensor assay.

### Mono-Cycle and Dual-Cycle NaSRED Protocols

Mono-cycle protocol was used in all the studies. Cultured food samples were also analyzed using dual-cycle NasRED protocol to assess the impact of food matrices on Stx2 detection sensitivity and resolution.

### Mono-Cycle Protocol

Samples were processed as previously described in our prior study^37^. Briefly, each sample was centrifuged at 1200 g for 5 minutes, incubated at room temperature for 20 minutes to allow AuNP aggregation, and vortex agitated at 1950 rpm for 5 seconds to selectively resuspend free AuNPs but not the clustered AuNPs. Immediately after processing, samples were analyzed using the portable electronic detector (PED) to obtain optical extinction signals for quantitative assessment of Stx2 concentrations.

### Dual-Cycle Protocol

To improve signal resolution at mid-range Stx2 concentrations (pg/mL), a dual-cycle aggregation protocol was developed. In the first cycle, samples were subjected to centrifugation at 1600 g for 5 minutes, incubated at room temperature for 5 minutes and vortex agitated at 3000 rpm for 10 seconds to fully resuspend AuNPs. In the second cycle, the same sample was centrifuged again at 1200 g for 10 minutes, followed by incubation at room temperature for 25 minutes to allow enhanced cluster formation and a final vortex agitation step at 2100 rpm for 5 seconds to achieve optimal resuspension of free AuNPs while preserving aggregated clusters. Samples containing functionalized AuNPs but not Stx2 (negative control, NC) were used to optimize the vortexing and incubation conditions to ensure assay specificity and reproducibility.

### DARPin-RED Data Processing

The data analysis for NaSRED consisted of signal acquisition, calibration, and normalization to ensure sensitive detection of Stx2 in biological and food matrices. First, target-containing samples were analyzed using the PED, alongside a negative control (NC) consisting of 18 μL of functionalized AuNPs mixed with 6 μL of the respective 4% biological media or food matrix in PBSD.

Each sample (18 μL AuNP sensing solution and 6 μL target solution, total 24 μL) and the NC tube were subjected to messurements at five random orientations in the tube holder to account for variations in tube positioning and matrix-induced optical effects, such as turbidity and color interference. The measured signals from the photodiode sensor were averaged, collected into a .csv datasheet, and saved to a local folder through an automated Python script.

The initial transmission signal at a given target concentration, T_0_(C), was the average of the five measurements before any processing steps (centrifugation, incubation, and vortexing). After the sensing protocol was completed, another five measurements were taken, yielding a post-processing average signal, T_t_(C). The difference in optical extinction (T_D_) was determined as follows:

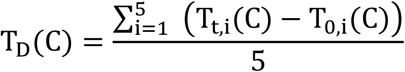

T_D_ value is proportional to Stx2-catalyzed AuNP clustering/sedimentation, which is influenced by Stx2 concentration. To ensure the reliability of the protocol based on our recent NasRED mechanism study^37^, the signal difference of negative control samples (T_D_(NC)) needs to be below a threshold criterion (*i.e.* T_D_(NC) <6% T_0_(NC)) to ensure that most AuNPs remain in suspension in the absence of Stx2 and the observed T_D_(C) signal is due to Stx2-catalyzed AuNP clustering.

To account for batch-to-batch variations in AuNP concentration and matrix-dependent signal fluctuations, all collected signals were further normalized to a range of 0 to 1, using a positive control (PC) sample as the reference, where all AuNPs were fully precipitated. The normalized sensing signal was calculated as:

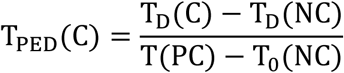

Given that all mediums in the study such as PBS, serum, and food matrices at 4% are clear, the PC reference was generated by mixing 18 μL of 1 × PBS dilution buffer with 6μL of the such matrices, representing the fully precipitated AuNP state.

For sensing in whole blood (WB), due to complex interactions between AuNPs and blood components, the PC reference was prepared using 18 μL of co-binder sensing solution mixed with 6 μL of 4% WB in PBTD with a high Stx2 concentration (C_H_= 100 pM). The PC signal was then defined as:

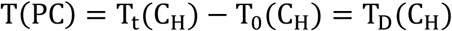

Additionally, measurement uncertainties were calculated as the standard deviation (SD) of the five orientation-specific normalized signals using:

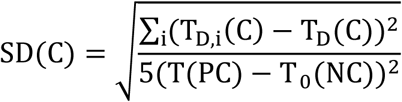

The final normalized NaSRED sensing signals, T_PED_ (C) ± SD(C), were plotted against Stx2 concentrations for each tested biological or food matrix samples to evaluate detection performance.

### Data Fitting, Lod, and LoQ Calculations

The normalized NaSRED sensing signals (T_PED_) were analyzed and fitted using Origin 2024b software (OriginLab, USA). To ensure precision, the orthogonal distance regression (ODR) algorithm was employed, incorporating both data values and error weights in the fitting calculations.

A biphasic dose-response model was used for data fitting using the following equation:

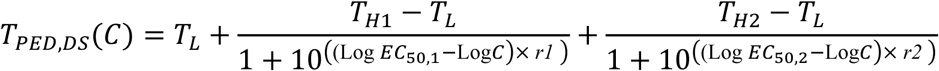

Where 𝑇_𝐻1_ and 𝑇_𝐻2_ represent the high signals for two different concentration phases, 𝑇_𝐿_ represents the lowest signal, obtained from the negative control (NC) or lowest analyte concentrations, 𝐸𝐸𝐶𝐶_50,1_ and 𝐸𝐸𝐶𝐶_50,2_ are the half-maximal effective concentrations for the respective phases of the biphasic response, *r1* and *r2* define the steepness of each segment of the curve. The dynamic range of the sensor was optimized by maximizing signal contrast (Δ𝑇*_PED_* = 𝑇_𝐻_ − 𝑇_𝐿_) and minimizing the steepness (r) to ensure a broad detection range. Fitting parameters were adjusted to optimize statistical metrics, ensuring Reduced Chi-square approaching 1, R-squared coefficient of determination (COD) close to 1, and Reduced sum of squares (RSS) approaching 0.

The LoD and LoQ were determined based on statistical differentiation from the NC signal, using standard deviation-based thresholds^52,53^. The LoD was defined as the lowest concentration at which the PED signal could be distinguished from the NC sample by 1.645 and 3 times the combined standard deviation of the NC and lowest concentration sample (*C_Min_*), calculated as:

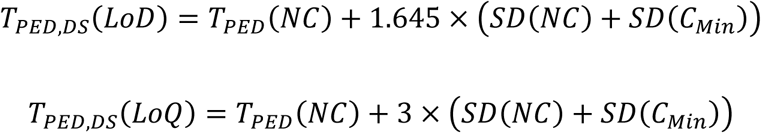

### Signal to Noise Ratio (SNR)

The mean signal-to-noise ratio (SNR) was calculated by averaging individual SNR values (𝑆𝑆𝑁𝑁𝑆𝑆(𝑐𝑐_𝑀𝑀_)) across all tested concentrations (𝑛𝑛) in the detection range:

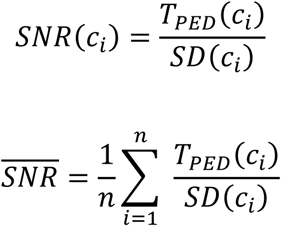

## Data Availability

All data produced in the present study are available upon reasonable request to the corresponding author

## Author Contributions

S.M. designed the experiments, performed data analysis, and wrote the original draft. C.W. and Z.C. conceived the idea of the DARPin-RED biosensor and supervised the research. Y.C. assisted in experiments and developed the PED readout system. K.C. contributed to manuscript preparation and worked alongside Z.C. in discussions regarding DARPin selection and validation. B.T. expressed and purified the DARPins and Stx2a toxins used in the study under the supervision of Z.C.. X.H. provided the food samples and Stx2 subtypes and participated in research discussions. C.W. and Z.C. supervised the project, directed experimental design and data interpretation, and revised the manuscript. All authors participated in discussions and manuscript editing.

## Declaration of Generative AI and AI-Assisted Technologies in the Writing Process

After completing the manuscript revision, the authors used OpenAI’s ChatGPT-4o as a reference in selected sections of the manuscripts to enhance clarity. The authors thoroughly reviewed and edited the content as necessary and take full responsibility for the final published version.

## Competing Interests Statement

C.W. and S.M. are co-founders of REDX Diagnostics, LLC. While the underlying technology may be subject to future commercialization, the affiliation did not influence the reported research findings. The remaining authors declare no competing interests.

## ACKNOWLEDGMENT

This project was supported in part by the National Science Foundation (NSF) under grant no. 1847324, U.S. Department of Agriculture AFRI 2022-67021-37013, and by the National Institute of Health under grant no. R21AI169098, <u>R21AI186134,</u> and DP2GM149552.

## Supplementary information

**Figure S1.**
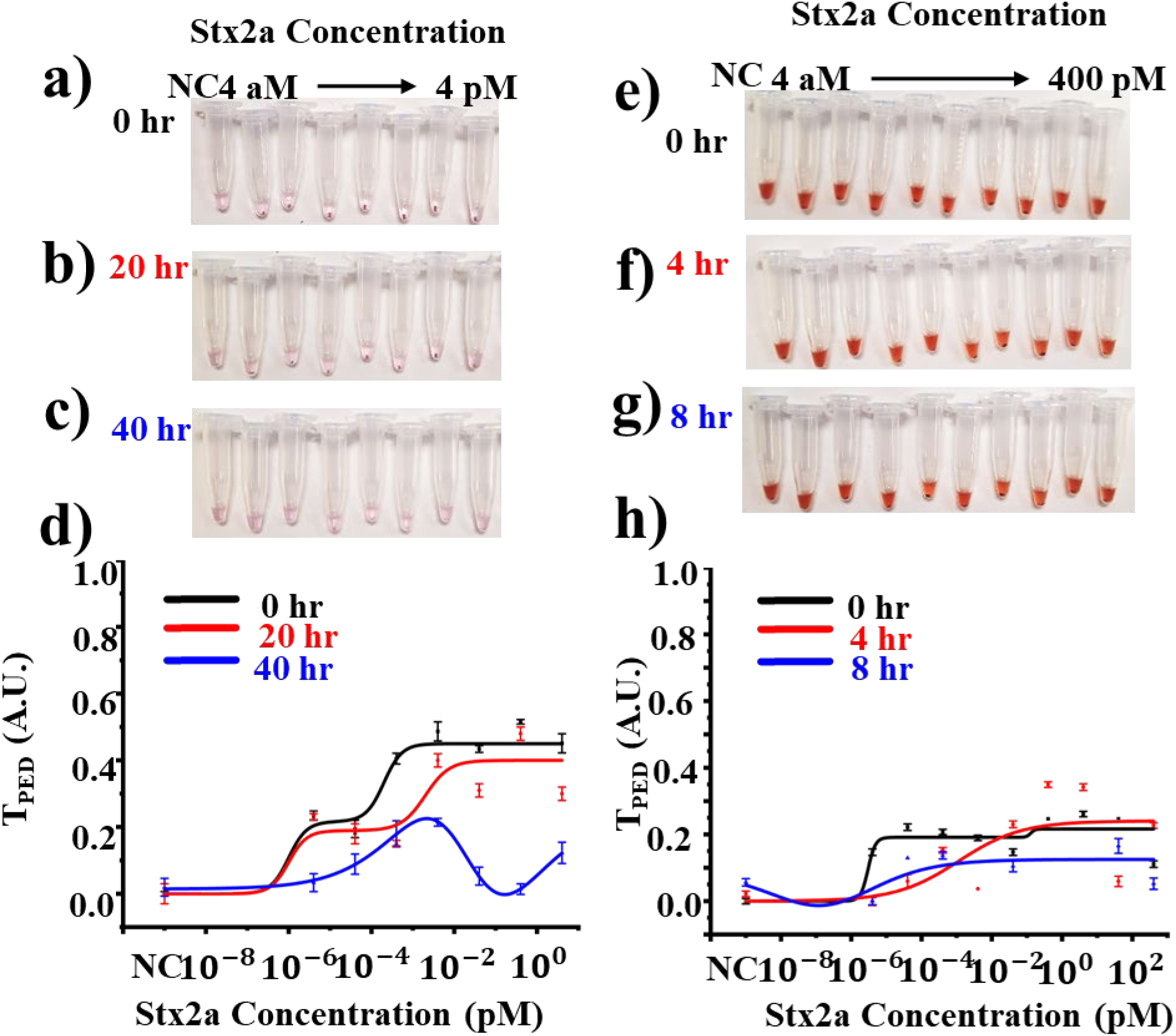
HuSAP effect analysis on signal performance in blood related samples. (a-c) Optical pictures of the testing tubes right before readout for Stx2a recombinant toxin spiked in 100% HPS stored at room temperature for (a) immediate detection after spike (0 hours), (b) 20 hours incubation, and (c) 40 hours of room temperature incubation. (d) extracted PED signals plotted against the Stx2 concentration in 100% HPS indicating signal response for 0 hours (solid black line), 20 hours (solid red line) and 40 Hours (solid blue line) indicating a shift in sensor response after 20 hours and diminished detection after 40 hours of incubation. (e-f) Optical image of testing tubes right before detection for Stx2a recombinant toxin spiked in 20 % whole blood and stored at room temperature for (e) 0 hours (immediate detection after spik), (f) 4 hours, and 8 hours. (h) extracted PED signals for Stx2 a spiked samples in 20 % whole blood after 0 hours (immediate detection, solid black line), 4 hours (solid red line) and 8 hours (solid blue line) after room temperature incubation. The sensing curve shifted to higher concentrations after 4 hours of incubation in 20 % blood and signals diminished after 8 hours of incubation indicating severe matrix and HuSAP interference in undiluted matrices.

**Figure S2.**
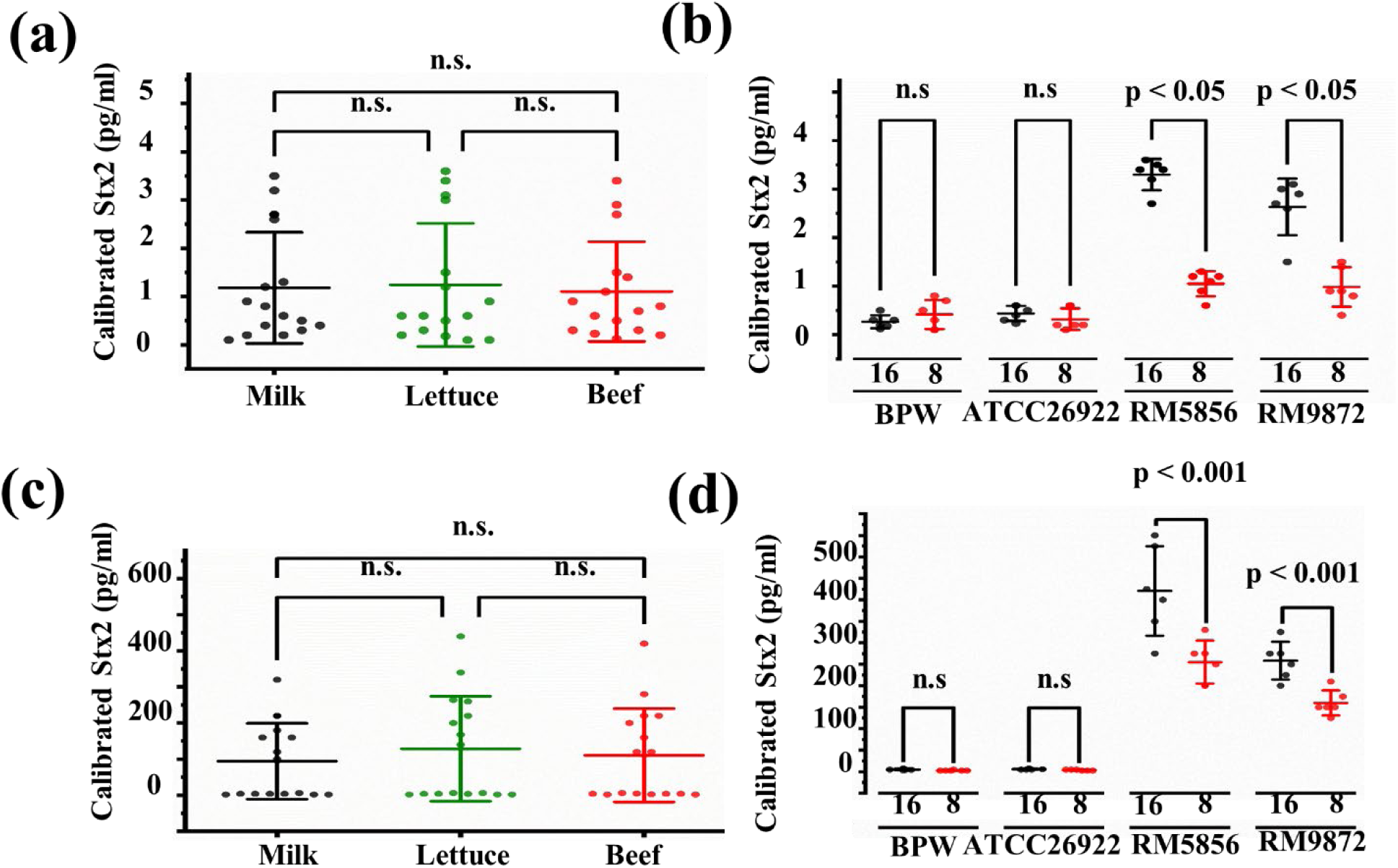
Statistical analysis of sensor performance for Stx2 quantification in STEC inoculated food samples. (a) Tukey’s two-way ANOVA of matrix effects, showing no significant difference (p > 0.05) between sensor responses, indicating minimal matrix interference in mono-cycle protocol. (b) Two-way ANOVA comparing bacterial strain and incubation time, showing significant differences (p < 0.05) between E. coli strains RM 5856 and RM 9872, demonstrating sensitivity to incubation duration in mono-cycle protocol. (c) Tukey’s two-way ANOVA of matrix effects, indicating no significant difference (p > 0.05) in sensor response across different matrices in dual cycle protocol. (d) Two-way ANOVA comparing bacterial strain and incubation time, revealing highly significant differences (p < 0.001) between Stx2-producing E. coli (RM 5856 and RM 9872) at different incubation times in dual cycle protocol.

## Notes

### Competing Interest Statement

The authors have declared no competing interest.

